# Prospective and External Evaluation of a Machine Learning Model to Predict In-Hospital Mortality

**DOI:** 10.1101/19000133

**Authors:** Nathan Brajer, Brian Cozzi, Michael Gao, Marshall Nichols, Mike Revoir, Suresh Balu, Joseph Futoma, Jonathan Bae, Noppon Setji, Adrian Hernandez, Mark Sendak

**Author notes:** Contact Information for Corresponding Author: Name: Nathan Brajer, Mailing address: 2806 Alderman Lane, Durham, NC 27705. Authors contributed equally to this work (Nathan Brajer, Brian Cozzi, Adrian Hernandez, Mark Sendak).

## Abstract

The ability to accurately predict in-hospital mortality for patients at the time of admission could improve clinical and operational decision-making and outcomes. Few machine learning models have been developed to predict in-hospital death that are both broadly applicable to all adult patients across a health system and readily implementable, and, to the best of our knowledge, none have been implemented, evaluated prospectively, or externally validated.

The primary objective of this study was to prospectively and externally validate a machine learning model that predicts in-hospital mortality for all adult patients at the time of hospital admission. Model performance was quantified using the area under the receiver operating characteristic curve (AUROC) and area under the precision recall curve (AUPRC). Secondary objectives were to design the model using commonly available EHR data and accessible computational methods.

A total of 75,247 hospital admissions (median [IQR] age 59.5 [29.0] years; male [45.9%]) were included in the study. The in-hospital mortality rates for the training validation, retrospective validations at Hospitals A, B, and C, and prospective validation cohorts, respectively, were 3.0%, 2.7%, 1.8%, 2.1%, and 1.6%. The area under the receiver operating characteristic curves (AUROCs), respectively, were 0.87 (0.83-0.89), 0.85 (0.83-0.87), 0.89 (0.86 – 0.92), 0.84 (0.80-0.89), and 0.86 (0.83-0.90). The area under the precision recall curves (AUPRCs), respectively, were 0.29 (0.25-0.37), 0.17 (0.13-0.22), 0.22 (0.14-0.31), 0.13 (0.08-0.21), and 0.14 (0.09-0.21).

The results demonstrated accurate prediction of in-hospital mortality for adult patients at the time of admission. The data elements, methods, and patient selection make the model implementable at a system-level.

## Introduction

An average of 2% of patients admitted to US hospitals die during the inpatient admission^1^. Efforts to reduce preventable in-hospital mortality have focused on improving treatments and care delivery^2^, and efforts to reduce non-preventable mortality have focused on supporting patient preferences to die at home^3-5^ and attempting to reduce health care costs in the inpatient setting. Early identification of patients at high risk of in-hospital mortality may improve clinical and operational decision-making and improve outcomes for patients^6^.

Previously developed machine learning models that predict in-hospital mortality have not been implemented or evaluated prospectively^7-10^, and most have not been externally validated^7-9^. Prospectively evaluating the performance of machine learning models run on real-world, operational data is a crucial step towards integrating these models into the clinical setting and evaluating the impact on patient care. External validation on patient data from distinct geographic sites is needed to understand how models developed at one site can be safely and effectively implemented at other sites^11^.

Unfortunately, formidable barriers prevent prospective and external evaluation of machine learning models and their integration into clinical care. First, after developing a model on retrospective data significant investment is needed to integrate the model into a production electronic health record (EHR) system. Second, most in-hospital mortality models are disease or department-specific and only evaluated on local data, limiting the ability to scale the benefit that can be realized from a single model^12-17^. Third, many efforts to date use either proprietary data sets, or advanced, computationally intensive modelling methods^7^, which limit adoption due to the technical capabilities required for successful integration.

The primary aim of this study was to prospectively and externally validate a machine learning model that predicts in-hospital mortality at the time of hospital admission for adult patients. The model uses readily available EHR data and accessible computational methods, can be applied to all adult patients, and was designed to be integrated into clinical care to support workflows that address the needs of patients at high risk of mortality. We evaluate the model on retrospective data from three hospitals and prospectively on a technology platform integrated with the production EHR system.

## Methods

### Setting

This study was performed at Duke University Health System (DUHS), a quaternary, academic, three-site hospital system with 1,512 beds that had approximately 69,000 inpatient admissions in fiscal year 2018. It adopted an Epic Systems EHR system in 2013. This study was approved by the Duke University Health System Institutional Review Board, which waived the need for informed consent for the use of identifiable data.

### Model training and development

We trained the model using EHR data from a total of 43,180 hospitalizations representing 31,003 unique adult patients admitted to a quaternary academic hospital (Hospital A) from October 1, 2014 to December 31, 2015. The outcome label of in-hospital mortality was defined as a discharge disposition of “expired”. 195 model features were built using 57 EHR data elements, which included patient demographics (5), as well as labs (33), vitals (9), and medication administrations (10) that occurred between presentation to the hospital and the time of inpatient admission. Time of inpatient admission was defined as time of order placement to admit a patient. The labs and vitals were aggregated by taking the minimum, maximum, mean, and variance of the recorded values and the medications were aggregated by the count of each medication type. Missing values for a given variable were assigned a default value that allowed us to include observations with missing data and enabled the algorithm to gain signal from missingness itself^18^. The data elements and model features are provided in the supplementary materials (eFigure 1).

We randomly selected 75% of encounters for model training, and held out 25% for testing. Initial tests demonstrated superior performance of gradient boosting models compared to random forest and regression models, so we selected a Gradient Boosting model using Python (with the XGBoost package^18-19^) as our base model with parameters chosen using cross validation. We used the CalibratedClassifierCV^20^ in Scikit-learn^21^ with 10 cross validation folds to calibrate the output so the predictions would better correspond to observed probabilities.

For each iteration of the cross validation, the model fit the 9 training folds with the XGBoost model. This fitted model was then used to generate predictions for the holdout fold. These predictions were then calibrated using Isotonic Regression. From this procedure, we trained 10 models, each consisting of an XGBoost prediction step and an Isotonic Regression step. To generate a prediction on new patient, we took the arithmetic mean of the output of these 10 models.

### Performance Metrics

To assess model discrimination, the receiver operating characteristic (ROC) and the precision recall (PR) curves were plotted and the respective areas under the curves (AUROC and AUPRC) were obtained. The AUPRC was included because it is particularly informative when evaluating binary classifiers on imbalanced data sets, where the number of negative outcomes significantly outweigh the number of positive outcomes^22,23^. Model calibration was assessed by comparing the predicted and empirical probability curves. 95% confidence intervals for the AUROC and AUPRC were obtained by bootstrapping the observations of the test data. Model output was plotted against observed mortality rates to assess calibration. Additionally, to assess performance in a manner that is meaningful to operational stakeholders, sensitivity, specificity, positive predictive value (PPV), and number of true and false alarms per day were generated for various risk thresholds. Lastly, model performance was evaluated on different patient subpopulations, segmented by information missingness, age, race, gender, admission types, and admission sources. Sensitivity, specificity, and PPV were obtained for each subpopulation using a set risk-threshold that resulted in a PPV of 20% across the entire population.

### Retrospective validations

A separate cohort of 16,122 hospitalizations representing 13,094 unique adult patients admitted to this same quaternary academic hospital (Hospital A) from March 1, 2018 to August 31, 2018 was used to assess temporal generalizability. Two separate cohorts from different community-based hospitals (Hospital B and Hospital C) were used to assess external generalizability. Hospital B had a cohort of 6,586 hospitalizations representing 5,613 unique adult patients and Hospital C had a cohort of 4,086 hospitalizations representing 3,428 unique adult patients. Both external cohorts consisted of admissions between March 1, 2018 and August 31, 2018. The AUROC and AUPRC for these three validations are reported.

### Prospective validation

The model was integrated into the production EHR system and prospectively validated at Hospital A, the quaternary academic hospital that the model was initially trained on. The prospective cohort consisted of 5,273 hospitalizations representing 4,525 unique adult patients admitted between February 14, 2019 and April 15, 2019. The model was run daily on a pipeline that automatically curates EHR data^24^ and the patient encounter identifier and risk scores were stored in a database over a 2-month period. Model output was not exposed to clinicians and was not used in clinical care during this silent period. At the end of the silent period, in-hospital mortality outcomes were obtained for each patient encounter. Model performance was assessed using methods identical to the retrospective validations. The AUROC and AUPRC for this prospective cohort was reported.

### Implementation

During the course of this study, we partnered with clinical and operational leaders to better understand how this model should be used in the hospital setting. As part of this work, we iteratively developed a simple user interface (eFigure 3 in the Supplement) showing model output using Apache Superset^25^, in addition to a workflow decision framework (eFigure 4 in the Supplement). Lastly, we developed a model facts sheet, similar to an over-the-counter drug label, that clearly communicates important information about the model to clinician end users (Figure 2).

## Results

We included a total of 75,258 hospitalizations in the training, retrospective validation, and prospective validation of the model. The overall percent of hospitalizations with in-hospital mortality was 2.7% (n = 2021). The hospital and encounter characteristics are summarized by training and testing cohorts in Table 1.

**Table 1.** Characteristics of hospitalizations in training and test data sets

|  |  |  |  |  |  |
| --- | --- | --- | --- | --- | --- |
|  | **Training data** | **Retrospective** | **Retrospective** | **Retrospective** | **Prospective** |
|  | **Hospital A** | **Hospital A** | **Hospital B** | **Hospital C** | **Hospital A** |
|  | **2014-2015** | **2018** | **2018** | **2018** | **2019** |
| Admissions |  |  |  |  |  |
| Hospitalizations, no. | 43180 | 16122 | 6586 | 4086 | 5273 |
| Length of stay, median (days) | 3.82 | 3.81 | 3.0 | 3.1 | 3.52 |
| Demographics |  |  |  |  |  |
| Age in years, median (IQR) | 58.6 (29.0) | 60.1 (29.1) | 59.0 (32.3) | 66.0 (21.0) | 59.7 (31.0) |
| Male sex, no. (%) | 20359 (47.1%) | 7375 (45.7%) | 2574 (39.1%) | 1906 (46.6%) | 2347 (44.5%) |
| Discharge disposition |  |  |  |  |  |
| Home, no. (%) | 35645 (82.5%) | 13231 (82.1%) | 5215 (79.2%) | 3144 (76.9%) | 4537 (86.0%) |
| Skilled nursing facility, no. (%) | 3937 (9.1%) | 1576 (9.8%) | 736 (11.2%) | 541 (13.2%) | 487 (9.23%) |
| Another healthcare facility, no. (%) | 589 (1.4%) | 201 (1.2%) | 279 (4.2%) | 105 (2.6%) | 49 (0.89%) |
| Rehabilitation, no. (%) | 564 (1.3%) | 204 (1.3%) | 90 (1.4%) | 87 (2.1%) | 22 (.042%) |
| Other, no. (%) | 159 (0.4%) | 112 (0.7%) | 58 (0.9%) | 34 (0.8%) | 16 (0.30%) |
| Expired, no. (%) | 1298 (3.0%) | 435 (2.7%) | 119 (1.8%) | 84 (2.1%) | 84 (1.6%) |
| Primary outcome |  |  |  |  |  |
| In-hospital deaths, no. (%) | 1298 (3.0%) | 435 (2.7%) | 119 (1.8%) | 84 (2.1%) | 84 (1.6%) |

Table 2 summarizes the prediction accuracy by evaluation method, site, and time period. For the retrospective validations, the AUROC was 0.87 for the 25% held-out test portion of the original training data set (retrospective evaluation, Hospital A, 2014-2015), 0.84 for a temporal validation cohort (retrospective evaluation, Hospital A, 2018), 0.89 for an external temporal and geographic validation cohort (retrospective evaluation, Hospital B, 2018), and 0.84 for an additional external temporal and geographic validation cohort (retrospective evaluation, Hospital C, 2018). For the prospective validation, the AUROC was 0.86 (prospective validation, Hospital A, 2019). The AUPRC, which compares sensitivity (recall) and positive predictive value (precision) and is more dependent on prevalence of the outcome, was lower across the 2018 retrospective and 2019 prospective validations compared to the earlier 2014-2015 retrospective validation. The prevalence of the outcome decreased from 3.0% in the 2014-2015 training data set compared to 1.6% in the prospective validation cohort.

**Table 2.**
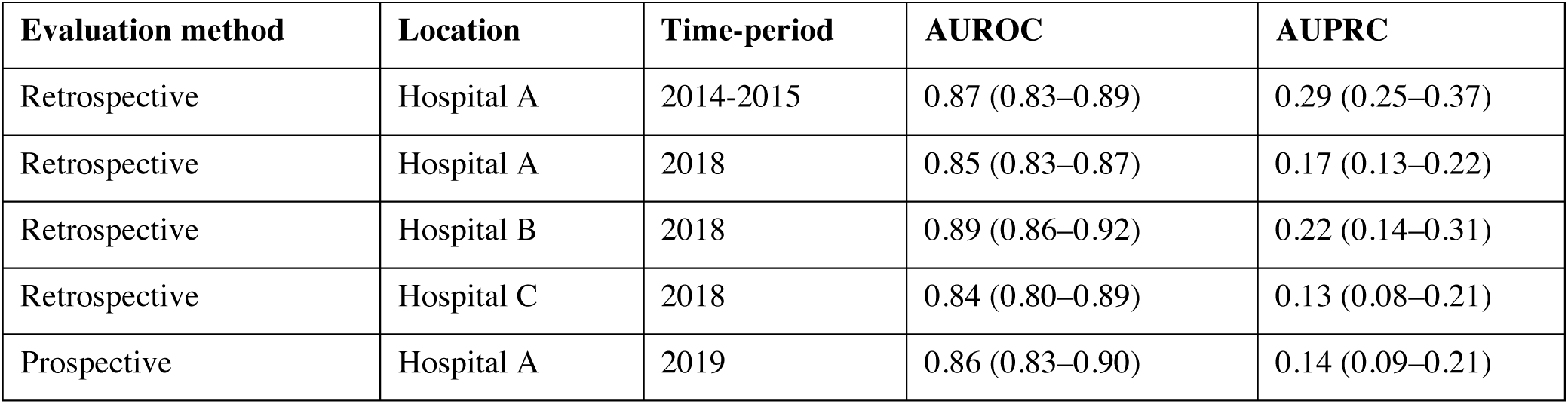
Prediction accuracy by evaluation method, location, and time-period

In Figure 1, the ROC and PR curves for the training test set demonstrate that the model performs well across risk threshold values, and the calibration plot demonstrates that mortality risk predicted by the model approximates observed mortality rates. The ROC and PR curves for the other evaluations are shown in the supplementary materials (eFigure 2).

**Figure 1.**
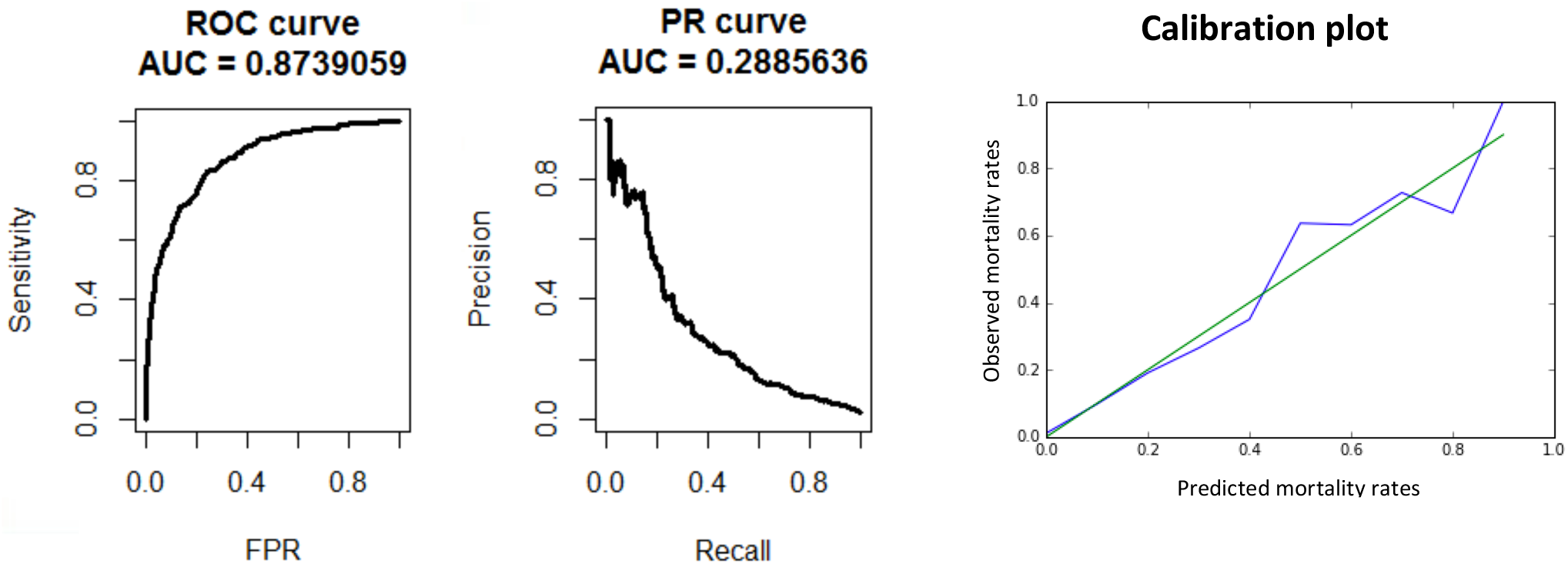
Receiver operating characteristic curve, precision recall curve, and calibration plot for 2014-2015 test set

Table 3 shows metrics that are most relevant to operational stakeholders in the 2014-2015 test set and assumes a theoretical admission volume of 100 patients per day. At this volume, alerts triggered at or above a threshold of 0.04 would be 61% sensitive, 91% specific, and result in 11.9 alerts per day. Of all fired alerts, 1.5 would be true alerts and 10.4 would be false alerts. At this risk threshold, the work-up-to-detection ratio is 7.9. As shown in Table 3, these metrics change significantly with different risk thresholds.

**Table 3.** Operational performance metrics for various model thresholds for the entire 2014-2015 test set, assuming a theoretical admission volume of 100 patients per day

|  |  |  |  |  |  |  |
| --- | --- | --- | --- | --- | --- | --- |
| Threshold | Sensitivity | Specificity | PPV | Total Alerts/Day | False Alerts/Day | True Alerts/Day |
| 0.01 | 0.88 | 0.66 | 0.05 | 39.9 | 37.8 | 2.1 |
| 0.02 | 0.76 | 0.81 | 0.08 | 23.3 | 21.5 | 1.8 |
| 0.03 | 0.68 | 0.88 | 0.11 | 15.3 | 13.6 | 1.7 |
| 0.04 | 0.61 | 0.91 | 0.12 | 11.9 | 10.4 | 1.5 |
| 0.05 | 0.57 | 0.93 | 0.15 | 9.1 | 7.7 | 1.4 |
| 0.06 | 0.54 | 0.95 | 0.18 | 7.4 | 6.1 | 1.3 |
| 0.07 | 0.52 | 0.95 | 0.19 | 6.5 | 5.3 | 1.3 |
| 0.08 | 0.50 | 0.96 | 0.21 | 5.8 | 4.5 | 1.2 |
| 0.09 | 0.48 | 0.96 | 0.22 | 5.2 | 4.1 | 1.2 |
| 0.10 | 0.44 | 0.97 | 0.22 | 4.8 | 3.7 | 1.1 |
| 0.11 | 0.43 | 0.97 | 0.24 | 4.4 | 3.4 | 1.0 |
| 0.12 | 0.41 | 0.97 | 0.24 | 4.1 | 3.1 | 1.0 |
| 0.13 | 0.39 | 0.98 | 0.26 | 3.7 | 2.7 | 1.0 |
| 0.14 | 0.39 | 0.98 | 0.27 | 3.5 | 2.6 | 0.9 |
| 0.15 | 0.36 | 0.98 | 0.27 | 3.2 | 2.3 | 0.9 |
| 0.16 | 0.35 | 0.98 | 0.28 | 3.1 | 2.2 | 0.9 |
| 0.17 | 0.34 | 0.98 | 0.30 | 2.8 | 2.0 | 0.8 |
| 0.18 | 0.33 | 0.98 | 0.32 | 2.6 | 1.7 | 0.8 |
| 0.19 | 0.31 | 0.99 | 0.32 | 2.4 | 1.6 | 0.8 |
| 0.20 | 0.29 | 0.99 | 0.33 | 2.2 | 1.5 | 0.7 |
| 0.21 | 0.28 | 0.99 | 0.33 | 2.0 | 1.4 | 0.7 |
| 0.22 | 0.28 | 0.99 | 0.35 | 1.9 | 1.3 | 0.7 |
| 0.23 | 0.27 | 0.99 | 0.36 | 1.8 | 1.2 | 0.7 |
| 0.24 | 0.26 | 0.99 | 0.38 | 1.7 | 1.1 | 0.6 |
| 0.25 | 0.26 | 0.99 | 0.41 | 1.5 | 0.9 | 0.6 |

eTable 1 shows model performance across patient subpopulations in the 2014-2015 test set. The risk threshold was selected to achieve a positive predictive value (PPV) of 20% across the entire 2014-2015 retrospective test set. The threshold of 0.075 results in an operationally manageable level of 6 total alerts per day (n=100 admissions per day), with 5 false alerts and 1 true alert. Model sensitivity, specificity, and positive predictive value (PPV) varied across patient subpopulations segmented by information missingness, age, race, gender, admission types, and admission sources.

eFigure 3 in the supplementary materials shows an Apache Superset dashboard that was developed to display patient risk scores to clinicians to support the development of workflows utilizing model output. The patient list can be filtered by time range to include patients admitted in the last day, last week, last month, and last year, and also includes an option for searching for a specific patient MRN. The dashboard is built off a curated database that automatically updates daily from the EHR reporting database. eFigure 4 in the supplementary materials shows the framework used to develop clinical workflows to be supported by model output. Figure 2 shows a model facts sheet, which summarizes important model information in a style similar to an over-the-counter drug label. The fact sheet is intended to guide use of the model by clinical end users who may not be familiar with the inner workings of the machine learning model.

**Figure 2.**
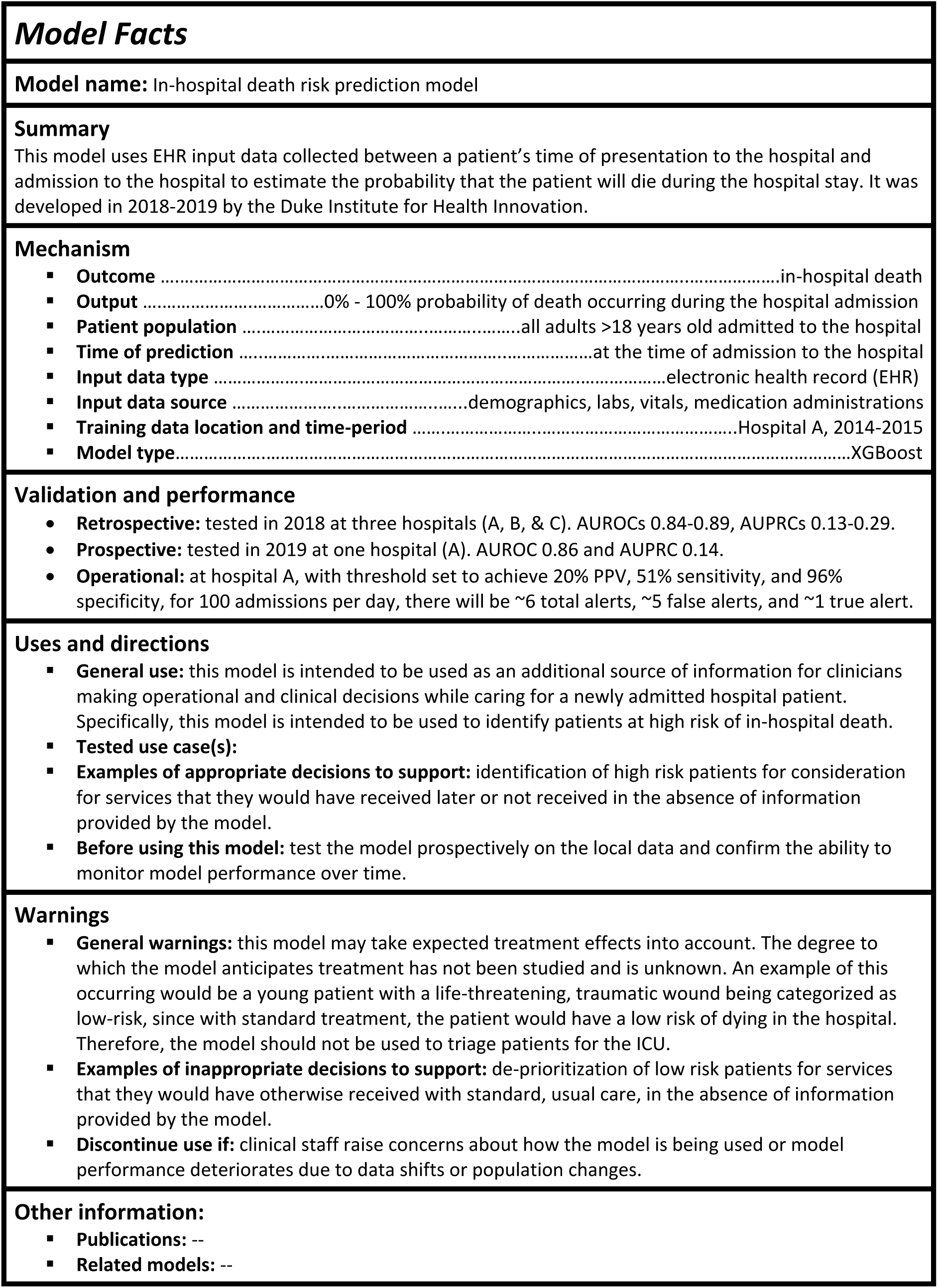
Model facts sheet to communicate important model information to clinical partners

## Discussion

We developed a machine learning model to predict in-hospital mortality at the time of admission for all adult patients using data that is readily accessible in the EHR. We retrospectively evaluated the model to assess temporal and external geographic generalizability and we prospectively evaluated model performance using a data pipeline that is integrated with our operational EHR. The model uses off-the-shelf software packages^18-20^ and was successfully integrated into a production EHR and can be similarly integrated at other sites.

To make the model implementable at a system-level, the model was trained on all adult patients using data elements commonly available across sites. The model only uses information from the current encounter without pre-hospital information, meaning that model outputs are accurate for patients who present for the first time to a health care setting. There are previously published models that predict inpatient mortality at the time of admission^7-10^. Rajkomar et al use deep learning in combination with tokenized data processing built on FHIR to train site specific mortality models. The model ingests all data available for a patient up until the time of prediction and achieves an AUROC of 0.90. The model does not require manual curation of features, which we have shown can cost more than $200,000 per application^26^. However, implementing this type of model requires potentially more substantial investment, including mapping all EHR data to FHIR resources and over 200,000 hours of graphical processing unit training time^27^. To our knowledge, the model has not yet been integrated into an operational EHR and evaluated prospectively. Tabak et al use logistic regression and conversion to an integer-based score (ALaRMS) to achieve an AUROC of 0.87. The model ingests demographic data and initial lab results across 23 numeric labs to predict inpatient mortality at the time of admission. The model was developed using data from 70 hospitals in the United States and has been externally validated by an independent team in Switzerland^10^. In addition to externally validating ALaRMS, Nakas et al developed decision tree and neural network models that ingested the same demographic and lab result inputs and both models achieved an AUROCs of 0.91. Finally, Schwartz et all use penalized logistic regression to develop a model that ingests demographics, initial lab results during a hospital encounter in addition to pre-hospital comorbidities to predict inpatient mortality^9^. The model achieves an AUROC of 0.857 and has not been externally or prospectively validated. To our knowledge, our model is the first to be integrated into an operational EHR and prospectively validated.

Prospective validation on operational data is an important first step in assessing the real-world performance of machine learning models. Ensuring that a model continues to perform well during a silent period sets the stage for integration into clinical workflows and evaluation of clinical and operational impact. Unfortunately, few machine learning models have been evaluated prospectively using real-world EHR data^28^, although there have been several recent validations of medical imaging models in the real world^29-31^. Our successful prospective validation is the first described in the literature for inpatient mortality prediction using a production EHR. The results in Table 2 show that model performance prospectively (AUROC = 0.86) almost exactly matched model performance retrospectively across the original test set (AUROC = 0.87) and the 2018 cohort (AUROC = 0.85). The AUPRC in the prospective cohort (0.14) was lower the retrospective AUPRC at the same hospital in 2014-2015 (0.29), which in part may have been due to the 47% reduction in prevalence of in-hospital mortality in the prospective cohort compared to the retrospective cohort (1.6% vs 3.0%). Overall, these results demonstrate that model performance remained stable when evaluated prospectively on operational data.

External validation results suggest stability in model performance when run on data from non-training sites and outside of the training time period. The AUROC in hospitals B and C in 2018 (0.89, 0.84) are consistent with the AUROC for training hospital A in 2014-2015 and 2018 (0.87, 0.84). Model stability over time and across settings suggest the model may not need to be re-trained, updated, or replaced as frequently. Furthermore, the possibility that models trained on data from one academic hospital in a health system can perform well at community hospitals in the same system may enable health systems to scale the clinical benefit realized from a single model without needing to rebuild site-specific the models.

In addition to performing well prospectively and on external sites, for machine learning models to improve patient care, they must also change behavior^32^. This requires improving decisions, which in the hospital, are usually made by physicians. During our efforts to develop workflows for this model with our clinical partners, we utilized a framework of identifying important decisions, specifying the decision-maker, understanding the time point at which the decision is made, verifying that there is an opportunity to improve, and confirming that the improvement can be measured (eFigure 4 in the Supplement). The dashboard (eFigure 3 in the Supplement), which shows a list of patient risk scores generated on a daily basis, coupled with the model facts sheet (Figure 2) helped clinicians understand the types of decisions that the model should and should not be used to support. Ultimately, this aided the process of identifying use-cases that can be tested and measured.

There is growing consensus that machine learning model reporting needs to be more standardized for a clinical audience^33^ as well as more transparent to end users^34^. The model fact sheet displayed in Figure 2 is the first formal effort in health care to distill important facts about how a model is developed and how a model should be used into 1 page. The fact sheet has undergone several iterations incorporating feedback from internal stakeholders. The need for such an artifact emerged after conversations with end users about the various ways the model output could inform clinical decision making. One especially troubling use of the model is called out in the model fact sheet. The model should not be used to prioritize patients for admission to the intensive care unit (ICU) or to limit care delivery. The model is built to predict risk of mortality at any point in time during the encounter and patients may receive life-saving therapies during an encounter that decrease risk of inpatient mortality. A study from 2015 describes how asthma was identified by a machine learning model as protective against death for patients admitted to a hospital with pneumonia^35^. The reason was that patients with asthma were admitted directly to an ICU and received appropriate care that was protective against inpatient mortality. If the model were prospectively implemented and incorporated into clinical decision making at the time of admission, it is not hard to imagine patients with asthma being de-prioritized for ICU admission. The downstream consequences are significant. The model should only be used to identify high risk patients for consideration for services that they would have received later or not received at all in the absence of information provided by the model. The first use-case that we plan to evaluate is identification of patients for palliative care consultation. The model fact sheet includes sections that are currently empty that we hope to populate as the evidence base for the current model grows.

## Challenges and Limitations

Many challenges were encountered in prospectively evaluating the model. First and foremost, the technology infrastructure required to run models in real-time had to be built. Fortunately, at our institution, such an infrastructure to automatically curate EHR data was already in place, utilizing native functionality in addition to custom developed technologies^24^. Second, there were many differences in data element names between the retrospective training data and prospective, live EHR data. This was due to changes in the EHR data structure over time that are not necessarily visible to the clinical end-user. Because of this, extensive mapping of variable names from the old retrospective data to the new prospective data was required. Third, differences in software versions between the computational research environment in which the model was trained and the production environment where the model was run had to be reconciled.

Our study has several potential limitations. First, we excluded patient less than eighteen years of age, and therefore our model will not be able to be used on pediatric patients. Second, it is rare for institutions to possess the technology infrastructure required to deploy models in a clinical setting on live EHR data and the scalability of this technical challenge is not addressed by our methods. The approach is scalable and effort and investment would likely be required by sites interested in adopting a similar approach. Third, the prospective validation was a silent period and did not include any interventions. While we demonstrate that the model prediction accuracy is maintained prospectively, it is unknown whether these predictions will actually improve clinical or operational outcomes. Future work will evaluate the implementation of a clinical workflow. Fourth, although we demonstrate external generalizability to community hospitals 4 and 30 miles from a flagship academic hospital, additional research is required to assess generalizability to more distant regions. Fifth, based on related work by others predicting inpatient mortality^5^ as well as prior internal experience predicting sepsis^36^, further performance gains may be achievable using deep learning. Lastly, further research is needed to understand how the model output should be interpreted and used in the clinical setting. For example, this study does not elucidate to what extent the model has learned treatment effects, and without careful instruction for how to interpret model output, clinicians may underestimate in-hospital mortality risk for patients with dangerous conditions that would usually receive intensive treatment. The goal of our model facts sheet is to highlight the recommended use cases as well as warn against use cases that may limit access to needed therapies in the inpatient setting. We hope the model fact sheet serves as a template for other teams conducting related work.

## Conclusion

Taken together, the findings in this study provide encouraging support that machine learning models to predict in-hospital mortality can be implemented on live EHR data with prospective performance matching performance seen in retrospective evaluations of highly-curated research data sets. This silent period validation sets the stage for integration into clinical workflows. The benefit to cost ratio of developing and deploying models in clinical settings will continue to increase as commonly available EHR data elements are more effectively utilized, and opportunities to scale models externally are identified. Further research is required to understand how to best integrate these models into the clinical workflow, identify opportunities to scale, and quantify the impact on clinical and operational outcomes.

## Data Availability

This study used electronic health record data from Duke University Health System. The study was approved by the Duke University Health System Institutional Review Board, which waived the need for informed consent for the use of identifiable patient data.

